# Combined talimogene laherparepvec and binimetinib in patients with *NRAS*-mutated melanoma induces anti-tumor immunity

**DOI:** 10.64898/2026.03.09.26347765

**Authors:** Tony Yao, Rita E Chen, Melissa Yamada, J Russell Moore, Marissa Jimenez, Tammy Huang, Lynn Cornelius, George Ansstas, Naresha Saligrama, David Y Chen

**Affiliations:** Department of Medicine, Washington University School of Medicine, St. Louis, MO; Medical Scientist Training Program, Washington University School of Medicine, St. Louis, MO; Division of Dermatology, Washington University School of Medicine, St. Louis, MO; Division of Oncology, Washington University School of Medicine, St. Louis, MO; Department of Neurology, Washington University School of Medicine, St. Louis, MO; Pathology and Immunology, Washington University School of Medicine, St. Louis, MO; Bursky Center for Human Immunology and Immunotherapy, Washington University School of Medicine, St. Louis, MO; Hope Center for Neurological Disorders, Washington University School of Medicine, St. Louis, MO; Siteman Cancer Center, St. Louis, MO

## Abstract

Immune checkpoint blockade can produce long-lasting responses in patients with metastatic melanoma; notably, combined CTLA-4/PD-1 blockade has been associated with approximately 52% melanoma specific 10-year survival (1). Yet, nearly half of patients experience minimal clinical benefit, and intensified regimens come with substantial risk of severe immune-related toxicity. The precise determinants of immunotherapy response are incompletely defined, reflecting a complex interplay between tumor biology and host immunity. This is especially consequential for patients whose disease progresses on checkpoint blockade, for whom effective salvage options are limited. In a series of patients with *NRAS*-mutated melanoma refractory to checkpoint inhibitors, we found that intratumoral administration of talimogene laherparepvec (T-VEC) combined with MEK inhibitor binimetinib induced exceptional clinical responses by amplification of pre-existing T cell responses and induction of *de novo* tumor-reactive immunity.

## Introduction

T-VEC is an oncolytic immunotherapy derived from herpes simplex virus type 1 that has been modified to enhance tumor-selective viral replication, oncolysis, and local immune priming through expression of immunostimulatory factors. T-VEC is approved for intratumoral administration into unresectable melanoma (2), and real-world data suggest that it remains effective for localized disease in the setting of immunotherapy salvage, even after multiple prior systemic therapies (3). In practice, T-VEC is considered a local therapy since it has not been consistently shown to exhibit significant rates of non-injected lesion response, and it does not have a clear, additive benefit when combined with anti-PD1 checkpoint blockade therapy (4). However, we observed that T-VEC combined with binimetinib induced exceptional responses in a series of patients with *NRAS*-mutated melanoma, including those with non-injectable disease.

## Results

Our index patient was a woman in her early 70’s with Stage IIIC melanoma of the right ankle, initially treated with surgical resection followed by one year of adjuvant nivolumab. Seven months after finishing adjuvant therapy, she developed cutaneous recurrence. She was treated with two cycles of ipilimumab and nivolumab but developed grade 4 interstitial nephritis, prompting checkpoint blockade therapy cessation. Her cutaneous melanoma continued to progress, and on presentation to our practice (**Figure 1A**, Patient 1), we initiated T-VEC and obtained tumor molecular profiling, which identified an *NRAS*^Q61R^ mutation. Binimetinib was added prior to the fourth cycle of T-VEC, and combination therapy resulted in rapid tumor regression. T-VEC was discontinued after 10 cycles while binimetinib was held two months later due to rhabdomyolysis, and she continues to have an ongoing complete response on observation.

**Figure 1.**
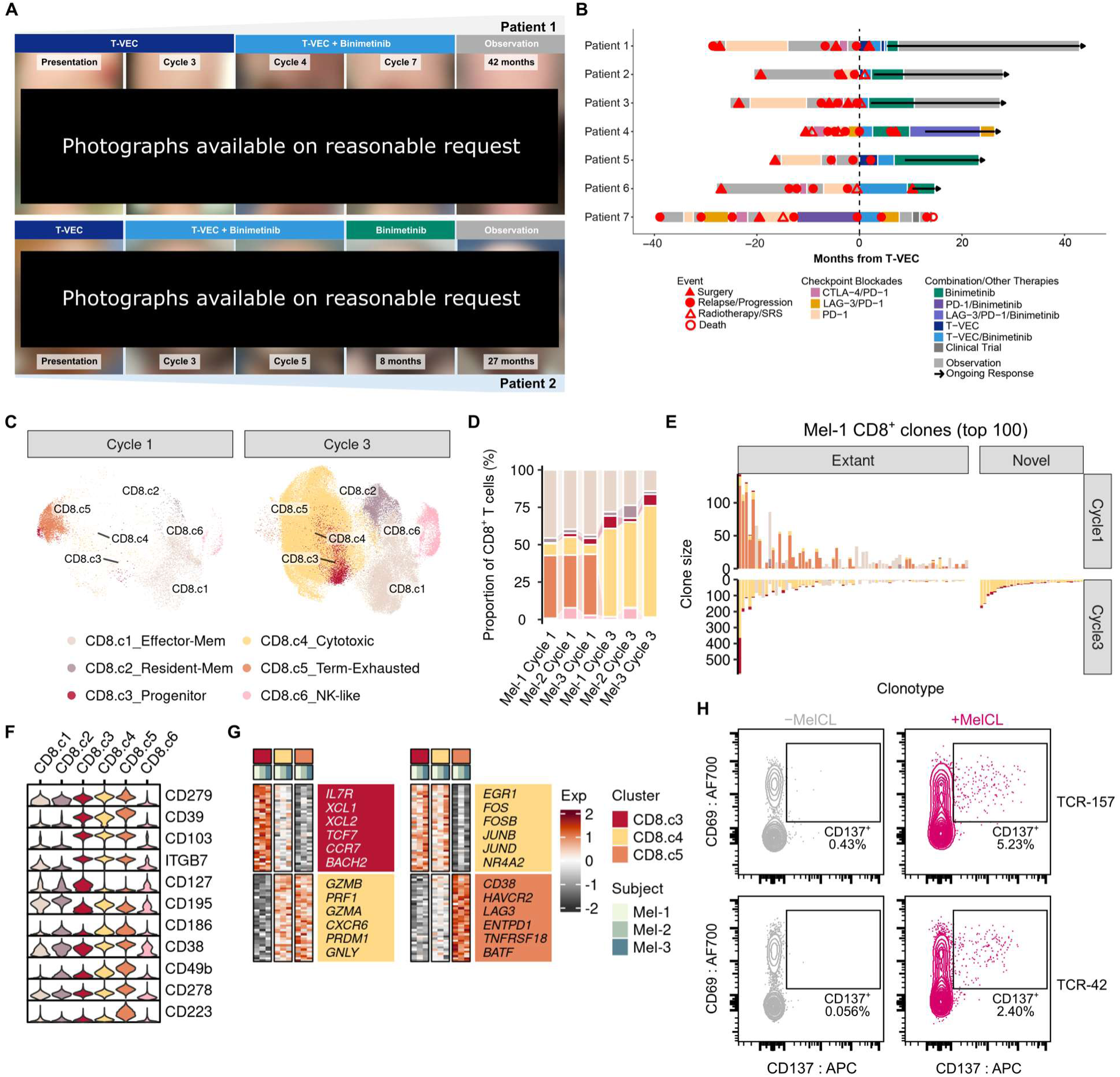
Revived anti-tumor CD8^+^ T cell immunity in complete responses to combined T-VEC and binimetinib therapy. (**A**) Cutaneous clinical response with combined T-VEC and binimetinib therapy in patients 1 and 2. (**B**) Clinical summary of all cases. (**C**) UMAPs of pre-and on-treatment intratumoral CD8^+^ T cells. Mem/Term, memory/terminal. (**D**) Stacked bar plots showing sample-wise phenotypic distribution of CD8^+^ T cells. (**E**) Stacked bar plots of phenotypic distribution among expanded CD8^+^ T cell clones from Mel-1. (**F**) Violin plots of surface protein expression across CD8^+^ T cell subsets. (**G**) Heatmaps of differentially expressed genes between progenitor, cytotoxic, and terminally exhausted CD8^+^ T cells. (**H**) Representative flow cytometry plots measuring activation of TCR-engineered effector CD8^+^ T cells with or without exposure to target autologous melanoma cells.

A consistent pattern emerged as additional patients with checkpoint-failed *NRAS*-mutated melanoma were treated using the same approach. In our observational cohort, 6 of 7 patients (85.7%) who received T-VEC and binimetinib (“combination”) therapy achieved a complete response (CR) compared to 2 of 7 (28.6%) patients with *NRAS*-mutated melanoma treated with T-VEC alone or combined with checkpoint blockade agents, which is in line with prior observations in unselected patients (3). Notably, the combination cohort had features associated with lower response rates to intratumoral therapy, including a higher frequency of distant (non-injected) disease (71.4% vs 14.3%) and brain metastases (42.9% vs 0%, **Table S1**). Responses in this group also occurred in the context of rapid progression prior to combination treatment in each case (**Figure 1B**), which is associated with worse T-VEC treatment outcomes (5). Importantly, these outcomes would not be anticipated from MEK inhibition alone, as binimetinib monotherapy rarely produces complete responses (1.5%) in *NRAS*-mutated melanoma patients (6).

To better understand the potential mechanisms underlying these responses, we conducted multiomic single-cell profiling on tumor biopsies from patients 2, 3, and 4 (Mel-1/2/3) collected before and during T-VEC and binimetinib combination therapy. We observed a marked local expansion of cytotoxic effector CD8^+^ T cells on therapy that replaced the existing terminally exhausted population (**Figure 1C-D**). This extensive phenotypic reconfiguration entailed consistent shifts across individual pre-treatment CD8^+^ clones from all patients (**Figure 1E, S1A-B**) and was not primarily attributable to clonal replacement as observed in checkpoint blockade (7). Both the cytotoxic and exhausted CD8^+^ T cell clusters, as well as a smaller progenitor pool with stem-like features, exhibited a CD39^+^ CD103^+^ tumor-reactive phenotype (**Figure 1F**) (8). Elevated surface LAG-3 and diminished expression of transcription factors which propagate T cell receptor (TCR) signaling distinguished the exhausted CD8^+^ state that was enriched before treatment (**Figure 1G, S1C**). We identified a group of clones with highly similar TCR sequences from patient 2, suggesting recurrent tumor antigen targeting (**Figure S1D**). To test tumor reactivity, we expressed two TCRs from this group in primary CD8^+^ T cells and co-cultured these with an autologous melanoma cell line derived from the pre-treatment tumor specimen (**Figure S1E**). We found that both a pre-existing (TCR-42) and a treatment-emergent clone (TCR-157) from this group demonstrated tumor reactivity (**Figure 1H-I**).

Intratumoral CD4^+^ T cells following combination therapy were enriched for a CXCR6^+^ Th1 effector subset distinct from the follicular helper (Tfh) phenotype associated with tumor-reactive CD4^+^ T cells (**Figure S2A-D**) (9). Pre-existing clones tended to retain their Tfh or Treg phenotype with expansion on treatment, while the Th1 state was largely restricted to newly emerging clonotypes (**Figure S2E**). Within expanded CD4^+^ clones, a minor fraction of cells could be found in a stem-like progenitor state (**Figure S2F**), and the majority exhibited an exclusive bias toward either the Tfh or Th1 phenotype (**Figure S2G**). No consistent changes in macrophage or DC subsets were found across treatment timepoints, but we observed an expansion of B and IgG1^+^ antibody-secreting cells, as well as upregulated IFN-inducible genes among blood endothelial cells at cycle 3 (**Figure S3**).

## Discussion

Evidence is emerging that the inability of T-VEC to induce systemic anti-tumor effects is due to antiviral immunity dominating T-VEC responses rather than development of anti-tumor reactivity (10). Our findings suggest that the addition of the binimetinib to T-VEC for *NRAS*-mutated melanoma may amplify and broaden anti-tumor responses, leading to exceptional clinical responses. Moreover, we have observed unexpected responses in melanoma patients with driver mutations in *HRAS*, *NF1*, and *GNAQ*, indicating that exceptional responses to combined T-VEC and binimetinib may extend beyond *NRAS*-mutated melanoma. This highlights the intriguing possibility that other tumors with MAP kinase pathway driver mutations may benefit from this combination therapy. Analysis of tumor biopsies during combination therapy revealed a renewal of existing CD8^+^ and recruitment of novel CD4^+^ Th1 clones, diverging from reported intratumoral T cell dynamics following checkpoint blockade (7, 9) and indicating orthogonal mechanisms of immune activation. Whether the observed influx of CD4^+^ Th1 cells reflects responses toward tumor or viral antigens, and provide help for priming of tumor-reactive CD8^+^ T cells (11), are critical open questions. These hypotheses should be formally tested, but we believe that the striking clinical responses compel us to report these results so that other patients might benefit from our experience.

## Data Availability

All data produced in the present study are available upon reasonable request to the authors.

## Acknowledgements

DC received support from K08 CA237727, MJ from the Two Sister’s Foundation. This work is supported in part by funding from the Alvin J. Siteman Cancer Center Catalyst Award and Department of Neurology start-up funds to NS. TY is supported by Canadian Institutes of Health Research Doctoral Foreign Study Award.

## SUPPLEMENTAL INFORMATION

### CASE DESCRIPTIONS

#### Patient 1

A woman in her late 60’s presented with concurrent superficial spreading melanoma, Breslow thickness 1.1mm, on the right ankle and superficial spreading melanoma, Breslow thickness 0.2mm, on the back. Three weeks later, both lesions were excised with sentinel lymph node biopsy revealing 4/8 examined nodes positive for melanoma, and staging imaging, including brain MRI, were negative for metastatic disease. She underwent adjuvant nivolumab therapy for one year. Seven months later, she developed a right ankle in-transit melanoma, negative for BRAF^V600E^ mutation, which was excised 12 weeks later. Three weeks following excision, she initiated ipilimumab plus nivolumab checkpoint blockade therapy, which was complicated by grade 4 acute interstitial nephritis causing treatment cessation after cycle 2. Follow-up PET-CT demonstrated metastatic right external iliac chain disease, and she was offered, but declined, palliative chemotherapy. She then presented to our multidisciplinary clinic where we performed a biopsy of a right shin lesion that we sent for next generation sequencing (**Figure 1A, top**). She initiated T-VEC therapy at this visit, and after four cycles of T-VEC, her leg lesions continued to expand and become increasingly symptomatic. Dermatologic surgery was consulted for palliative debulking surgery for the largest, most symptomatic nodule. The day prior to debulk, she added binimetinib in combination with T-VEC after sequencing results were positive for a NRAS^Q61R^ mutation. Surprisingly, much of the debulked mass was necrotic with <10% viable tumor. After 2 months of combined T-VEC and binimetinib, she developed hematuria, and binimetinib was held for 3 weeks before re-initiating at a reduced dose. She continued with T-VEC for 10 total cycles, after which she developed edema and erosions at the injection sites, so T-VEC was discontinued. Less than two weeks after that, she developed rhabdomyolysis and binimetinib was also discontinued. Since then, she has been on clinical observation and continues to have no evidence of disease 42 months after starting T-VEC.

#### Patient 2

A man in his early 70’s was found by his dermatologist to have a 1.9mm Breslow thickness, non-ulcerated melanoma for which he underwent wide local excision with sentinel node biopsy with one of three nodes positive for melanoma, negative for BRAF V600E mutation. Staging, including brain imaging, did not reveal metastatic disease, so he elected clinical surveillance with imaging and circulating tumor DNA blood screening. Fourteen months later, he was found to have cutaneous metastatic melanoma on the scalp, which was excised, following which he initiated adjuvant nivolumab. At this time his tumor was sent for sequencing, revealing an NRAS Q61R mutation. He was found to have significant cutaneous progression of the scalp after two cycles of nivolumab (**Figure 1A, bottom**). CT head and neck revealed new nodules on the right posterior scalp and neck representing metastatic disease, and brain MRI revealed new right posterior temporal lobe and right superior occipital lobe lesions suspicious for metastatic disease, which prompted initiation of combined ipilimumab and nivolumab checkpoint blockade. Given continued cutaneous progression, ipilimumab and nivolumab were discontinued, and he was referred to our multidisciplinary clinic where we proceeded with T-VEC and binimetinib. One month after T-VEC initiation, he received gamma knife SRS to his brain metastases. He experienced rapid clinical response, and after 5 cycles of T-VEC, we performed biopsies revealing pathological complete response with tumoral melanosis, and T-VEC was discontinued. During this time, he was also noted to have an acneiform rash, a well-described cutaneous side effect of MEK inhibitors. He also developed dropped head syndrome, which is an uncommon side effect of MEK inhibitor; binimetinib was discontinued 6 months later, resulting on complete resolution of his cervical myopathy. He remains without evidence of disease on observation 27 months after starting T-VEC and binimetinib.

#### Patient 3

A man in his mid-30’s presented with a lesion on the right forearm with a 1.1mm Breslow thickness Spitzoid melanoma for which he underwent wide local excision and sentinel lymph node biopsy two months later with 1/1 lymph nodes positive for melanoma. He then underwent one year of adjuvant nivolumab therapy. Fourteen months later, he developed a recurrent melanoma in his right forearm scar for which he underwent wide local excision and sentinel lymph node biopsy where 0/4 nodes were positive. One month later, he developed an occipital metastatic melanoma lesion. PET-CT revealed a new, hypermetabolic right level 5 cervical lymph node suspicious for metastatic disease while brain MRI was negative for intracranial metastases. He underwent neoadjuvant ipilimumab plus nivolumab for 2 cycles followed by scalp lesion resection and right neck dissection, finding 2/69 nodes positive for melanoma. One month later, PET-CT revealed new FDG-avid nodules on his right forearm that were biopsied, revealing recurrent melanoma. Sequencing demonstrated *NRAS*^Q61K^ mutated melanoma, prompting binimetinib initiation followed by T-VEC the following week. Decreased nodularity in the right forearm was noted after four cycles of T-VEC, and biopsies revealed complete pathologic response. After 11 months of binimetinib, his left ventricular ejection fraction (LVEF) was noted to be 43%, and binimetinib was held. His LVEF improved to 51% after 1 month of binimetinib discontinuation. He is under observation now and has continued to show no evidence of recurrence for 27 months after starting T-VEC and binimetinib.

#### Patient 4

A woman in her early 60’s presented with a left lower extremity mass growing over 6 months. Punch biopsy revealed an ulcerated, 20mm Breslow thickness nodular melanoma. Staging brain MRI revealed a hemorrhagic right parieto-occipital brain mass, and both primary melanoma and brain lesions were surgically resected. After pathologic confirmation of brain metastatic melanoma, she underwent stereotactic radiosurgery (SRS) to an intact brain lesion and radiotherapy to the resection bed. She then completed four cycles of ipilimumab and nivolumab, but restaging PET-CT two weeks later revealed an early recurrence at the left lower extremity, which was resected three weeks later while on maintenance nivolumab. MRI two weeks following identified brain metastases for which she received additional SRS while on nivolumab. At this time, she switched to relatlimab in combination with nivolumab, but three months later, biopsy of the left lower leg demonstrated recurrent melanoma which was sent for sequencing, revealing an *NRAS*^Q61K^ mutation. She then presented to our multidisciplinary clinic and started binimetinib in combination with T-VEC. After six cycles of T-VEC, she achieved complete cutaneous response, and T-VEC was stopped. Three months later, she developed a left frontal lobe recurrence while on binimetinib for which she underwent surgical excision and SRS. Two months later, she restarted nivolumab and relatlimab in addition to binimetinib. Binimetinib was discontinued 13 months later, and she continues to have no evidence of progression at 26 months since T-VEC initiation.

#### Patient 5

A woman in her mid-50’s presented with a right second toe 1.8mm Breslow thickness nevoid acral melanoma without ulceration. She then underwent ray amputation, but a sentinel lymph node biopsy was not performed due to lack of tracer activity. CT imaging was negative for distant disease. She remained on surveillance for six years until she developed an in-transit melanoma on her right foot which was excised, and PET-CT was negative for residual or distant disease. However, a new nodule had developed over the excision scar a month later, prompting her to complete 7 cycles of single agent nivolumab over 8 months but stopped due to clinical evidence of recurrence along her right shin. Three months later, she began treatment with ipilimumab plus nivolumab but stopped after two cycles when she developed grade 4 immune mediated colitis, which required treatment with infliximab and a high dose prednisone taper. Three months after discontinuing immunotherapy, a PET-CT demonstrated the presence of at least five FDG-avid lesions on the patient’s right shin consistent with progressive disease. A month afterwards, she initiated T-VEC therapy and completed 5 cycles in 3 months with some systemic side effects, including fever and malaise. Imaging demonstrated disease progression with development of a new hypermetabolic subcutaneous nodule on her right shin and increased size and metabolic activity of injected lesions. She then started binimetinib for NRAS^Q61R^ mutated disease and completed six additional, concurrent cycles of T-VEC over the next 4 months, achieving a complete response. During this time, binimetinib was held for less than a month due to elevations in her hepatic enzyme tests. She also developed an acneiform eruption in response to binimetinib, which was treated with doxycycline. Restaging PET-CT after seven total months of T-VEC injections revealed minimal residual FDG avidity along the right anterior tibia. Biopsy demonstrated changes suggestive of treatment effect. She continues to have no new or progressive disease while still on binimetinib 23 months after starting T-VEC.

#### Patient 6

A woman in her mid-40’s presented with a 1.1mm Breslow thickness, ulcerated melanoma of the right forearm that had been increasing in size for over a year. One month later, she underwent wide local excision with sentinel lymph node biopsy where 0/3 lymph nodes were positive for melanoma, and the melanoma was found to be negative for a *BRAF* mutation and positive for a *NRAS*^Q61K^ mutation. Subsequent imaging, including a brain MRI, revealed no evidence of metastatic disease. A year later, the patient presented with pain and purulent drainage from a lesion in her right axilla. She underwent a right axillary lymph node biopsy, which revealed metastatic melanoma. Two weeks later, a brain MRI identified a left occipital lesion, and a PET-CT showed a right axillary necrotic lymph node but no other sites of uptake. She then started ipilimumab and nivolumab therapy. completing three cycles but held further therapy due to hepatopathy requiring prolonged prednisone therapy. PET-CT two months later showed decreased metabolic activity in the previously identified right axillary lymph node but identified a new site of uptake in the right femur. She then started single agent nivolumab and completed six cycles when a follow-up PET-CT demonstrated disease progression and a brain MRI revealed a new left cerebellar lesion. Two months later, she underwent stereotactic radiosurgery of her brain lesions and began treatment with T-VEC injections combined with binimetinib. After completing 16 cycles of T-VEC, imaging studies suggested decreased right axillary uptake and no evidence of distant disease. So, she underwent surgical resection of her right axillary lymphadenopathy, but no viable tumor was identified, achieving a complete response which continues to endure 14 months after starting T-VEC therapy.

#### Patient 7

A woman in her early-70’s with a distant history of cutaneous melanoma (> 20 years prior) on the right arm and right lateral lower leg presented with a right inguinal metastatic melanoma diagnosed by core biopsy. A right, superficial inguinal node dissection revealed 2/18 lymph nodes positive for metastatic disease, and PET-CT three months later showed no evidence of recurrent or residual disease. However, 6 months afterwards, a repeat PET-CT demonstrated a metastatic lesion in the subcutaneous tissue of the anterior right thigh, which was excised. PET-CT around 17 months later revealed a pulmonary nodule and left adrenal gland lesion, both highly suspicious for metastatic disease, but the patient initially elected surveillance. She opted to begin treatment 5 months later with single agent nivolumab. After 3 months, CT imaging demonstrated disease progression, so she was transitioned to nivolumab and relatlimab. After 5 months, she switched to ipilimumab and nivolumab due to disease progression, completed 4 cycles, and underwent a left adrenalectomy. At this time molecular profiling revealed *NRAS*^Q61R^ mutation. While on maintenance nivolumab, around 7 months later, imaging identified a new nodule within the left adrenalectomy bed suspicious for recurrent disease, so she was started on binimetinib in combination with nivolumab. After a year, imaging demonstrated right inguinal nodal progression, so T-VEC was initiated for the right inguinal metastatic disease combined with binimetinib and nivolumab. She completed 9 cycles of T-VEC over the course of 4 months when CT imaging demonstrated disease progression in the chest in addition to new hepatic and gallbladder lesions suspicious for metastases. She discontinued T-VEC and completed 4 cycles of nivolumab and relatlimab but experienced disease progression. She then participated in a clinical trial in which she received the CD137 agonist CTX-471 at 0.1 mg/kg every 2 weeks and pembrolizumab every 6 weeks. She discontinued the trial about three months later when CT imaging confirmed disease progression. She then began treatment on a clinical trial with the BTLA antagonist TAB004 and toripalimab, completing two cycles of treatment before presenting with a left cerebellar hemorrhagic metastasis, resulting in her death.

In summary, this series of patients with *NRAS*-mutated melanoma has experienced unexpectedly high rates of complete response to combination T-VEC and binimetinib. In one case with progressive disease, the patient had progression while treated with binimetinib prior to initiating T-VEC, suggesting that tumor sensitivity to binimetinib is a required feature for combined agent responses and implicates mechanisms more broad than augmented infectivity of virus or immune responses previously described (12). The combination with binimetinib may also be consequential and distinguishing from other experience with MEK inhibitors combined with T-VEC (13).

### METHODS

#### Case selection and response criteria

This single institution retrospective cohort study was conducted in accordance with Washington University School of Medicine Human Research Protections Office (HRPO) approved protocols 201409103 and 202309151, which include informed consent for analysis of patient data and tissue samples and clinical photography. Patients eligible for this study were aged ≥18 years who had confirmed *NRAS* mutations by next generation sequencing, had received three or more doses of T-VEC therapy, and completed T-VEC-based therapy at Washington University. Eligible patients started T-VEC based therapy between January 1, 2018 and January 1, 2025 with a data cutoff date of December 15, 2025 to ensure adequate time for durable clinical response determination. We used investigator-assessed objective response (OR) for response determination as previously described (3). Briefly, progressive disease (PD) was determined by increasing number or growth of lesions necessitating a switch in therapy, while complete response (CR) was called by clinical or radiographic resolution of targetable lesions. All CRs in this study were confirmed by target lesion histopathology.

#### Tissue collection and processing

Tumor punch biopsies and blood were collected from patient participants who provided informed consent (HRPO #201409103) prior to T-VEC initiation and prior to cycle 3 of T-VEC. Tumor specimens were minced and digested in RPMI supplemented with 20% fetal bovine serum (FBS), collagenase type IV (Worthington CLS-4), and Benzonase (Sigma 70664) for 2 hours at 37°C, then manually dissociated through a 70-μm cell strainer to obtain single cell suspensions. PBMCs were isolated from whole blood by density gradient centrifugation with lymphocyte separation medium (Corning 25-072). Cells were viably cryopreserved in FBS containing 10% DMSO for downstream experiments.

#### Single cell sequencing

Cryopreserved pre-cycle 1 and pre-cycle 3 tumor biopsy cell suspensions from three patients (Mel-1/2/3) were thawed, stained, and sorted for cellular indexing of transcriptomes and epitopes (CITE-seq) with joint single cell T and B cell receptor sequencing (scTCR/BCR-seq). Thawed cells were stained with Zombie Green viability dye (BioLegend 423112), blocked with human TruStain FcX solution (BioLegend 422302), and labelled with oligo-conjugated TotalSeq-C (Supporting Data: “CITE-seq Antibodies”), respective hashing (BioLegend 394661, 394663, 394665, 394667, 394669, 394671), and BV421 anti-human CD45 (clone HI30, Biolegend 304032) and PE anti-human CD31 (clone WM59, BioLegend 303106) sorting antibodies in PBS with 0.5% bovine serum albumin (BSA). Live singlet CD45^+^ immune and CD45^−^ CD31^+^ endothelial cells were sorted using a BD FACSAria II, pooled, and resuspended in PBS with 0.04% BSA. Single cell partitioning, cDNA amplification, and generation of gene expression, V(D)J, and feature barcode libraries were performed according to the Chromium Next GEM Single Cell 5’ HT v2 protocol (10x Genomics CG000424). Libraries were sequenced on Illumina Novaseq X Plus systems by the Washington University Genome Technology Access Center.

#### Pre-processing of CITE-seq data

FASTQ files from gene expression and feature barcode libraries were respectively aligned to the GRCh38.p13 genome assembly (GENCODE v32) and a supplied TotalSeq-C feature reference, counted, and demultiplexed using Cell Ranger v7.2.0 multi (14) to generate count matrices for each GEM well. Feature-barcode matrices were imported and merged into a Seurat v5.0.0 object (15). Cell barcodes were filtered based on the number of detected gene expression features (300 < *nFeature_RNA* < 8,000), non-hashtag feature barcode UMIs (500 < *nCount_ADT* < 30,000), hashtag UMIs (200 < *nCount_HTO* < 20,000), percent of mitochondrial gene counts (*percent.mt* < 10%), and percent of ribosomal gene counts (*percent.rb* > 3%). Cells were assigned to their sample of origin (patient × timepoint) and hashtag-heterotypic doublets were excluded using the MULTIseqDemux function (quantile = 0.3) (16) on HTO counts. Feature barcode UMI counts were denoised and normalized using the DSBNormalize function (dsb v1.0.3) (17) with quantile clipping at 0.005 and 0.9999.

#### Dimensionality reduction, clustering, and visualization of CITE-seq data

Raw gene expression UMI counts were log-normalized and scaled. Principal component analysis (PCA) reductions were separately performed on the top 3,000 variable RNA features and DSB-normalized surface protein data. RNA assay data from different subjects were integrated using the Seurat reciprocal PCA method. A weighted-nearest neighbor graph was computed with the first 30 RNA PCs and first 20 DSB PCs, which was used as input for smart local moving clustering and UMAP visualization. This clustering procedure was applied first to categorize cells into broad lineages (CD8^+^ T, CD4^+^ T, NK, B/plasma, myeloid, endothelial cells), then iteratively on each subset to assign granular cell type labels. Differentially expressed genes between cell groupings were identified using Seurat implementation of Wilcoxon rank-sum tests and visualized with ComplexHeatmap v2.20.0 (18) as pseudobulk log-normalized data scaled by row. All other visualizations were generated with ggplot2 v3.5.1 or Seurat functions.

#### Processing and visualization of scTCR-seq data

FASTQ files from V(D)J libraries were assembled and annotated based on the prebuilt GRCh38_alts_ensembl-5.0.0 reference using Cell Ranger v7.2.0 multi to generate clonotype tables for each GEM well. High-confidence contig annotations (filtered_contig_annotations.csv) were appended as metadata to T cells in the CITE-seq dataset. For each patient, clones were defined as cells sharing TCRα/β chains with fully matched V(D)J gene segments and CDR3 nucleotide sequences, permitting α chain dropout only in cases where the β chain sequence is uniquely identifying. Clonotypes were categorized as extant if recovered from both pre-cycle 1 and cycle 3 timepoints and as novel if found only at cycle 3. Phenotypic distributions of expanded clones, ranked by cumulative frequency across timepoints, were visualized using ggplot2 v3.5.1.

#### Generation of primary melanoma cell line

In parallel to sorting immune and endothelial for single cell sequencing, live CD45^−^ CD31^−^ cells were sorted from pre-cycle 1 tumor biopsies to enrich melanoma cells for culture. Sorted cells were expanded and passaged in Opti-MEM (Gibco 51985034) supplemented with 5% FBS, 1 mM sodium pyruvate, 100 U/mL penicillin-streptomycin, 5 μg/mL insulin (Santa Cruz Biotechnology, sc-360248), and 5 ng/mL recombinant human EGF (Gibco, PHG0311). A primary melanoma cell line was derived from patient Mel-1 and aliquots of early-passage cells were cryopreserved. Whole-exome sequencing of genomic DNA from this cell line confirmed presence of the *NRAS*^Q61L^ driver mutation.

#### TCR engineering in primary T cells

Paired TCRα/β sequences were selected from expanded CD8^+^ T cell clones in Mel-1 and cloned as polycistronic TCRβ-P2A-TCRα inserts into the pLVX-EF1α-IRES-Puro lentiviral vector (Takara, 631988) by GenScript. Murine *Trac* and *Trbc1* sequences with stabilizing cysteine substitutions (19) were used for TCR construct constant regions. For viral packaging, Lenti-X 293T cells (Takara, 632180) were transfected using VSV-G Single Shots (Takara, 631282), then 48 h supernatants were collected, concentrated with Lenti-X Concentrator (Takara, 631231) per manufacturer’s protocols, and used without freezing.

Human CD8^+^ T cells were purified from PBMCs by magnetic negative selection (StemCell, 17953) and activated for 48 h in complete RPMI (cRPMI) containing 10% fetal bovine serum (FBS), 1 mM sodium pyruvate, 1× non-essential amino acids, and 100 U/mL penicillin-streptomycin supplemented with 25 ng/mL IL-7 (Miltenyi, 130-095-367), 50 ng/mL IL-15 (Miltenyi, 130-095-760), and TransAct anti-CD3/CD28 reagent (Miltenyi, 130-128-758). Immediately prior to transduction, endogenous TCR chains were disrupted in stimulated CD8^+^ T cells by CRISPR/Cas9 nucleofection (20). Ribonucleoprotein complexes (RNPs) were formed by mixing 180 pmol each of single guide RNA targeting human *TRAC* (IDT, TCAGGGTTCTGGATATCTGT) and *TRBC1/2* (IDT, CGTAGAACTGGACTTGACAG) with 60 pmol TrueCut Cas9 v2 (Invitrogen, A36499) and incubating for 10 min at room temperature. Up to 5 million stimulated CD8^+^ cells were resuspended in 20 μL supplemented P3 buffer (Lonza, V4XP-3032), mixed with RNPs, electroporated (pulse EH111), and rested in cRPMI for 15 min at 37°C. In parallel, lentiviral particles were loaded onto Retronectin-coated 24-wells (Takara, T100A) by centrifugation of concentrated lentiviral supernatants (2h, 2,000 g, 32°C). Rested CD8^+^ T cells were added to lentivirus-coated wells and spinoculated (30 min, 800 g, 32°C). Transduced CD8^+^ T cells were cultured in cRPMI containing IL-7 and IL-15 for 4 to 10 days before evaluation of transduction efficiency and tumor reactivity screening.

#### Flow cytometry

Cells were stained with Zombie NIR viability dye (BioLegend, 423106) in PBS for 15 min at 4°C, washed and resuspended in FACS buffer (0.5% BSA in PBS), blocked with human TruStain FcX for 10 min at 4°C, then stained with surface antibodies for 30 min at 4°C. For intracellular staining, fixation and permeabilization were performed using the eBioscience FoxP3/Transcription Factor Staining Buffer kit (Invitrogen, 00-5523-00) per manufacturer’s protocol. The following antibodies were used for flow cytometry: SV500 anti-human CD3 (clone SK7, BioLegend 344870), SB550 anti-human CD8 (clone SK1, BioLegend 344760), APC anti-human TCRα/β (clone IP26, BioLegend 306718), BV421 anti-human TCRα/β (clone IP26, BioLegend 306722), AF700 anti-human CD69 (clone FN50, BioLegend 310922), APC anti-human CD137 (clone S18014C, BioLegend 300806), APC anti-human SOX10 (clone SP267, Abcam ab310845), PE anti-human Melan-A (clone A103, Santa Cruz Biotechnology sc-20032), and BV711 anti-mouse TCRβ (clone H57-597, BioLegend 109243). Samples were acquired on a Cytek Aurora. FlowJo v10.10.0 was used for data analysis and visualization.

#### Tumor reactivity screening of TCR-engineered T cells

Primary melanoma cells from patient Mel-1 were thawed, expanded, and seeded into a flat-bottom 96-well plate (50,000 cells/well) in cRPMI. Melanoma cells were incubated for 4 h at 37°C and 5% CO2 to allow for attachment, after which TCR-engineered CD8^+^ T cells were added (250,000 cells/well) and co-cultured for 16 h. Wells without melanoma cells were included as negative controls. Following co-culture, upregulation of CD137 among TCR-engineered (human TCRα/β^−^ and mouse TCRβ^+^) CD8^+^ T cells was measured by flow cytometry as an indicator of reactivity toward target tumor cells.

## TABLE

**Table S1:** Comparison between T-VEC vs T-VEC + Binimetinib (TVEC + Bini) cohorts. *Some columns add up to >100% due to rounding

| Table S1: *NRAS* -mutated melanoma cohort |  |  |
| --- | --- | --- |
|  | T-VEC | T-VEC+Bini |
| **Sex** |  |  |
| Female | 4 (57.1%) | 5 (71.4%) |
| Male | 3 (42.9%) | 2 (28.6%) |
| **Age at T-VEC initiation** |  |  |
| Median (range) | 58 (50-82) | 64 (36-77) |
| **Tumor subtype** |  |  |
| Cutaneous | 6 (85.7%) | 5 (71.4%) |
| Acral | 1 (14.3%) | 1 (14.3%) |
| Mucosal | 0 (0%) | 1 (14.3%) |
| **History of distant metastases** |  |  |
| Yes | 1 (14.3%) | 5 (71.4%) |
| No | 6 (85.7%) | 2 (28.6%) |
| **Presence of non-injectable lesions at T-VEC start** |  |  |
| Yes | 1 (14.3%) | 5 (71.4%) |
| No | 6 (85.7%) | 2 (28.6%) |
| **Brain metastases** |  |  |
| Yes | 0 (0%) | 3 (42.9%) |
| No | 7 (100%) | 4 (57.1%) |
| **Prior lines of systemic therapy** |  |  |
| 0 | 3 (42.9%) | 0 (0%) |
| 1 | 3 (42.9%) | 0 (0%) |
| 2 | 1 (14.3%) | 6 (85.7%) |
| 3+ | 0 (0%) | 1 (14.3%) |
| **Prior immune checkpoint blockade** |  |  |
| Yes | 4 (57.1%) | 7 (100%) |
| No | 3 (42.9%) | 0 (0%) |
| **Concurrent systemic therapy** |  |  |
| Nivolumab | 1 (14.3%) | 0 (0%) |
| Pembrolizumab | 2 (28.6%) | 0 (0%) |
| Relatlimab/nivolumab | 1 (14.3%) | 0 (0%) |
| Binimetinib | 0 (0%) | 6 (85.7%) |
| Nivolumab + binimetinib | 0 (0%) | 1 (14.3%) |
| None | 3 (42.9%) | 0 (0%) |
| **Clinical response** |  |  |
| Complete (CR) | 2 (28.6%) | 6 (85.7%) |
| Partial (PR) | 2 (28.6%) | 0 (0%) |
| Stable disease (SD) | 0 (0%) | 0 (0%) |
| Progressive disease (PD) | 3 (42.9%) | 1 (14.3%) |

## SUPPLEMENTAL FIGURES

**Supplemental Figure 1.**
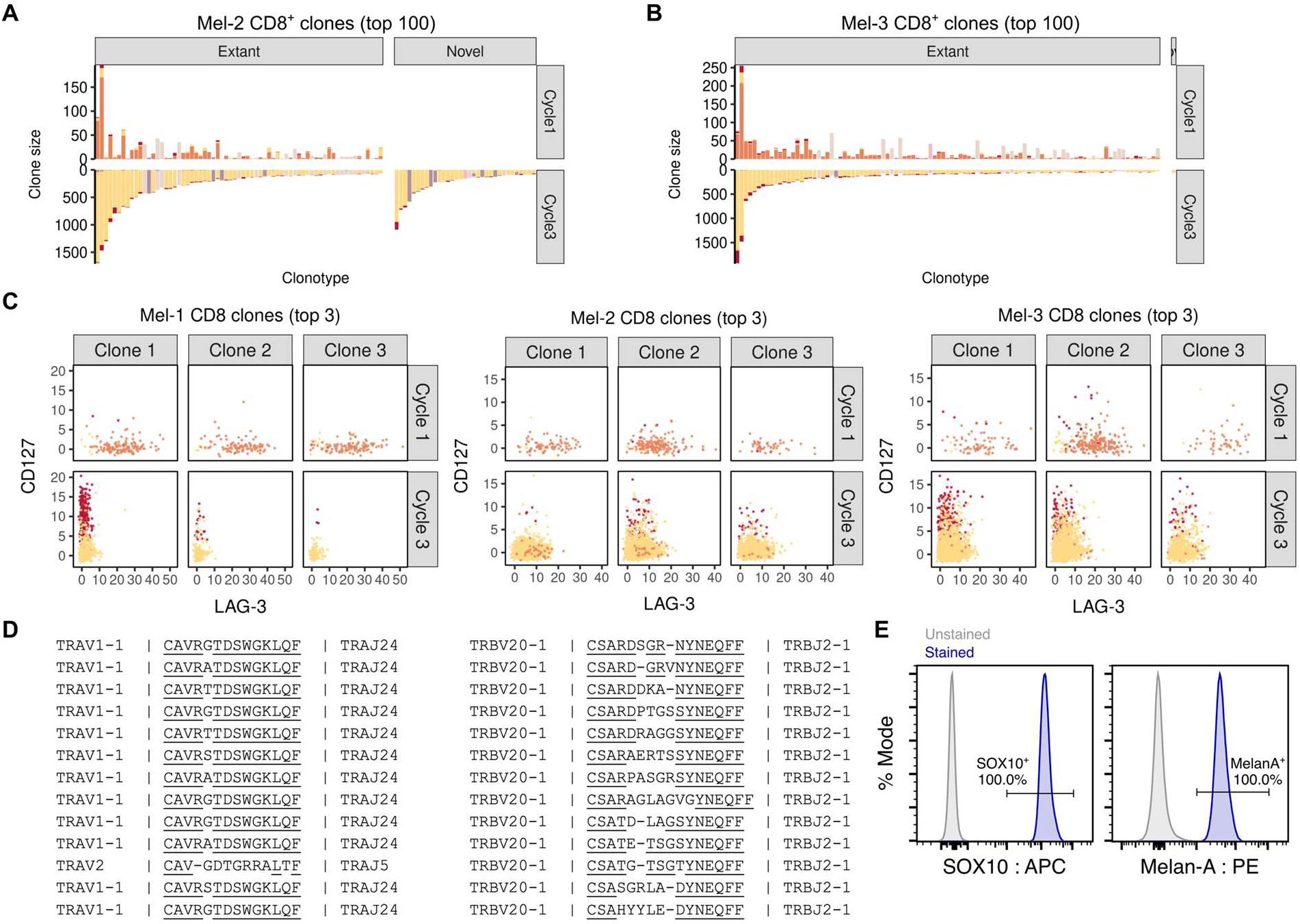
Pre-existing exhausted CD8^+^ T cell clones are found as cytotoxic effectors following combined T-VEC and binimetinib therapy. (**A-B**) Stacked bar plots depicting phenotypic distribution of expanded CD8^+^ T cell clones from Mel-2 (**A**) and Mel-3 (**B**). Shifts in surface phenotype across timepoints in selected representative CD8^+^ T cell clones. (**C**) Group of highly similar TCR sequences among Mel-1 CD8^+^ T cell clones. (**E**) Histograms displaying flow cytometric validation of melanoma marker expression in primary tumor cell line derived from the pre-treatment specimen of Mel-1.

**Supplemental Figure 2.**
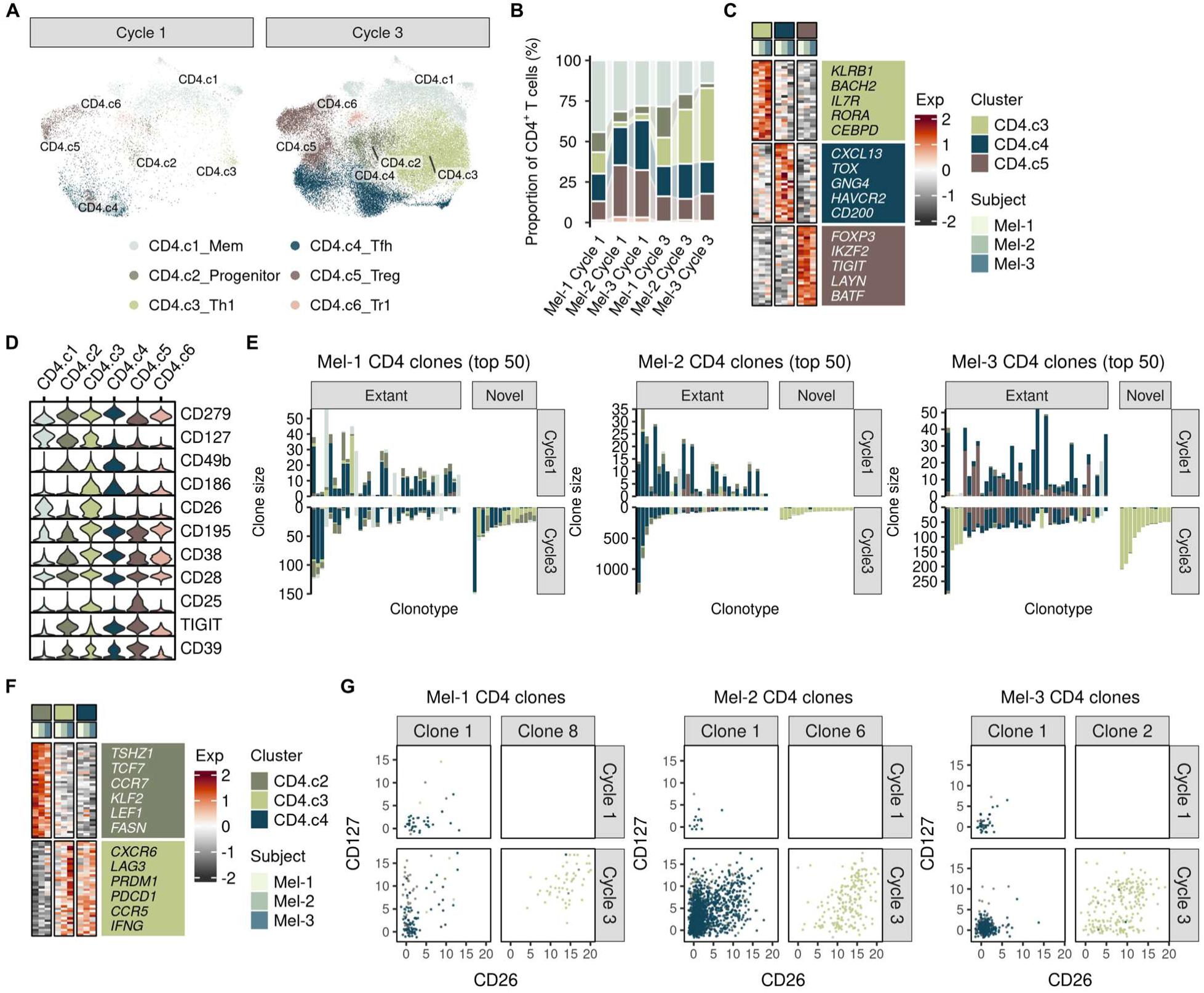
CD4^+^ T cell clonal dynamics following combined T-VEC and binimetinib therapy. (**A**) UMAPs of pre- and on-treatment intratumoral CD4^+^ T cells. Mem/Tfh/Tr1, memory/follicular helper/type 1 regulatory T cell. (**B**) Stacked bar plots depicting sample-wise phenotypic distribution of CD4^+^ T cells. (**C**) Heatmap of differentially expressed genes between Th1, Tfh, and Tr1 subsets. (**D**) Violin plots of surface protein expression among CD4^+^ T cell subsets. (**E**) Stacked bar plots depicting phenotypic distribution of expanded CD4^+^ T cell clones. (**F**) Heatmap of differentially expressed genes between progenitor and differentiated CD4^+^ T cell states. (**G**) Surface phenotype across timepoints of selected Tfh- and Th1-biased CD4^+^ T cell clones.

**Supplemental Figure 3.**
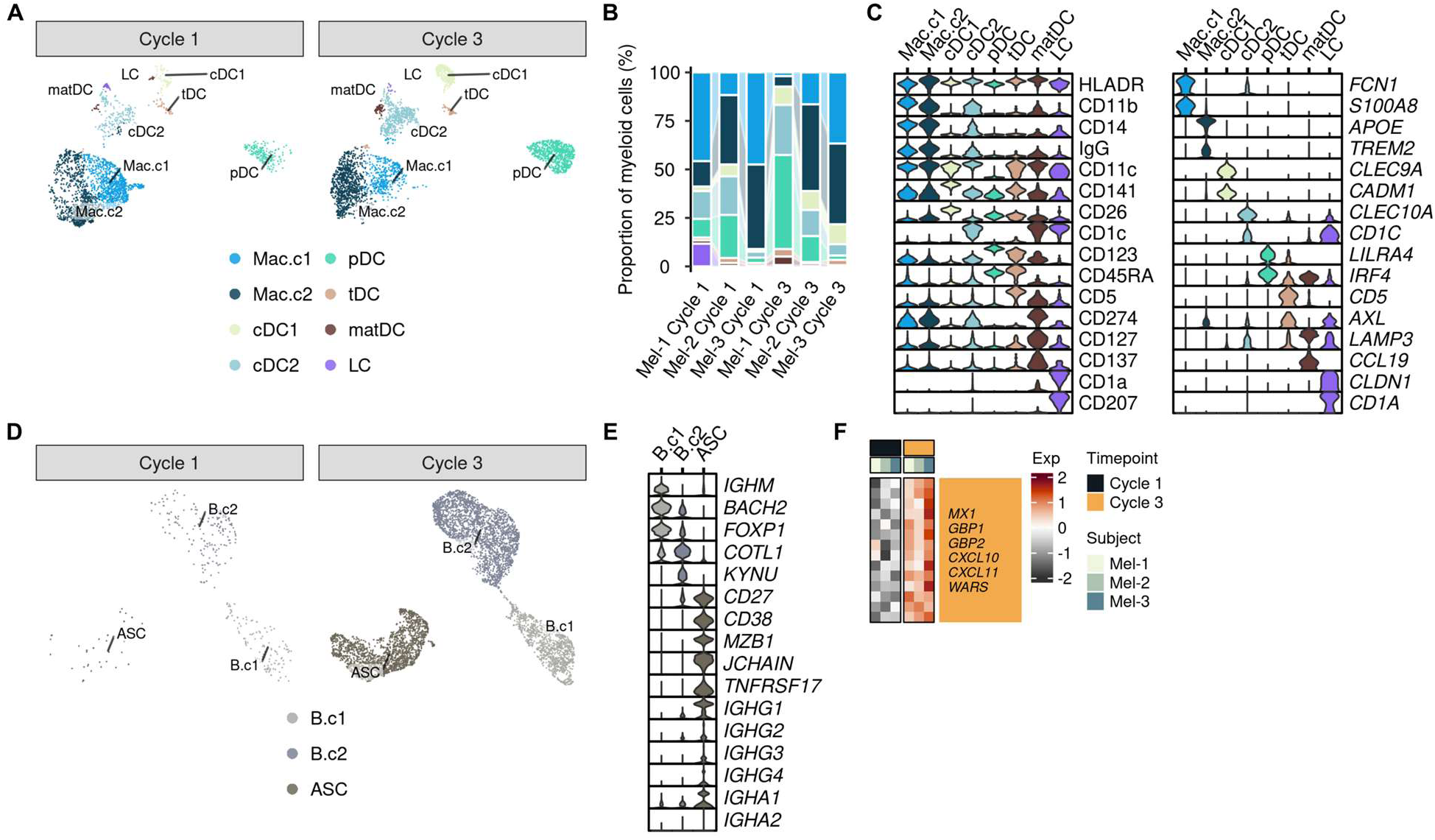
Changes in myeloid, B/plasma, and endothelial cell compartments following combined T-VEC and binimetinib therapy. (**A**) UMAPs of pre- and on-treatment intratumoral myeloid cells. Mac/cDC/pDC/tDC/matDC/LC, macrophage/conventional DC/plasmacytoid DC/transitional DC/mature DC/Langerhans cell. (**B**) Stacked bar plots depicting sample-wise phenotypic distribution of myeloid cells. (**C**) Violin plots of surface protein and gene expression among myeloid cell subsets. (**D**) UMAPs of pre- and on-treatment intratumoral B and antibody-secreting cells (ASCs). (**E**) Violin plots of gene expression among B cells and ASCs. (**F**) Heatmap of differentially expressed genes between timepoints among blood endothelial cells.

